# Performance of polygenic risk scores for cancer prediction in a racially diverse academic biobank

**DOI:** 10.1101/2021.05.13.21256833

**Authors:** Louise Wang, Heena Desai, Shefali S. Verma, Anh Le, Ryan Hausler, Anurag Verma, Renae Judy, Abigail Doucette, Peter E. Gabriel, Regeneron Genetics Center, Katherine L. Nathanson, Scott Damrauer, Danielle L. Mowery, Marylyn D. Ritchie, Rachel L. Kember, Kara N. Maxwell

## Abstract

**Purpose:** Genome-wide association studies (GWAS) have identified hundreds of single nucleotide polymorphisms (SNPs) significantly associated with several cancers, but the predictive ability of polygenic risk scores (PRS) is unclear, especially among non-Whites.

**Methods:** Genome-wide genotype data was available for 20,079 individuals enrolled in an academic biobank. PRS were derived from significant DNA variants for 15 cancers. We determined the discriminatory accuracy of each cancer-specific PRS in patients of genetically-determined African and European ancestry separately.

**Results:** Among individuals of European genetic ancestry, PRS for breast, colon, melanoma, and prostate were significantly associated with their respective cancers (OR 1.25-1.47). Among individuals of African genetic ancestry, PRS for breast, colon, and prostate were significantly associated with their respective cancers. The AUC of a model comprised of age, sex, and principal components was 0.617–0.709 and increased by 1-4% with the PRS in individuals of European genetic ancestry. In individuals of African genetic ancestry, AUC was overall higher in the model without PRS (0.740-0.811) but increased < 1% with the PRS in the majority of cancers.

**Conclusion:** PRS constructed from SNPs moderately increased discriminatory ability for cancer status in individuals of European but not African ancestry. Further large-scale studies are needed to identify ancestry-specific genetic factors in non-White populations to incorporate PRS into cancer risk assessment.

## INTRODUCTION

Cancer is the second leading cause of death nationally, with over 1.8 million new cancer cases and 600,000 deaths projected in the United States in 2020^1^. The development of risk-stratified models based on a combination of genetic and non-genetic risk factors allow personalized development of cancer prevention and surveillance strategies to reduce cancer morbidity and mortality^1-3^. For example, identification of rare genetic factors of high and moderate risk, such as *BRCA1/2* pathogenic variant status, are used to determine cancer surveillance and prevention strategies for breast cancer^4,5^. However, these factors account for less than 5% of breast cancer prevalence in large, population based studies^6,7^. Current strategies for breast cancer risk prediction for the majority of women rely upon models which incorporate non-genetic breast cancer risk factors such as age, race, family history^8^, estrogen-related factors^9^, BMI, and mammographic density^10^.

The incorporation of common genetic factors could vastly improve current risk prediction models for cancer. Genome-wide association studies (GWAS) have identified a large number of common genetic variants associated with multiple cancers^11^, but the risk association of each variant is small and impossible to individually incorporate into risk prediction models. Polygenic risk scores (PRS) are a weighted sum of multiple disease associated alleles to identify individuals at high risk for a specific disease or phenotype^12^. For example, PRS can stratify women above and below a lifetime breast cancer risk threshold of 20% which is used to justify the incorporation of breast MRI into cancer surveillance protocols^13^.

PRS improves the accuracy of breast cancer risk prediction models^2,4^; however, PRS is less well-studied in other cancer types^12^. In addition, as non-European patients are historically underrepresented in GWAS studies^14^, the weights derived from non-European GWAS and used for PRS models may not be appropriate for non-White patients^15^. Current commercial genetic tests using PRS are only available for individuals of European and/or Ashkenazi Jewish ancestry, which may worsen already existing cancer health disparities^16^.

We therefore sought to evaluate the discriminatory ability of PRS in predicting cancer risk for 15 cancers (bladder, breast, colorectal, endometrial, esophageal, glioma, lung, melanoma, oral cavity and pharynx, ovarian, pancreatic, prostate, renal, testicular, and thyroid) that had at least one GWAS study within the past 10 years in genetically determined African and European individuals using data from an academic biobank.

## MATERIALS AND METHODS

### Penn Medicine BioBank Cohort and genotyping

The Penn Medicine BioBank (PMBB) is a genomic and precision medicine cohort comprised of participants who actively consent for biospecimen collection and linkage of the their biospecimen to their electronic health record (EHR) data^17^. Participants were recruited between 2004-2020 to a University of Pennsylvania institutional review board approved study at the time of their medical appointment in the University of Pennsylvania Health System (n=60,232). As of December 2020, 20,079 unique participants (some samples were genotyped multiple times) had genome-wide DNA array-based genotyping using the Global Screening Array (GSA) chip in three batches: 1) 5,676 samples on Illumina GSA (Global Screening Array) V1 chip (SNPs = 700,078); 2) 2,972 samples on Illumina GSA V2 chip at Children’s Hospital of Philadelphia Center for Applied Genomics (SNPs = 759,993); 3) 16,940 samples on Illumina GSA V2 chip at Regeneron Genetics Center. All individuals who were recruited for the PMBB are patients of clinical practice sites of the University of Pennsylvania Health System. Appropriate consent was obtained from each participant regarding storage of biological specimens, genetic sequencing, access to all available EHR data and permission to recontact for future studies. The study was approved by the Institutional Review Board of the University of Pennsylvania.

### Genotyping quality control and genetic ancestry determination

Quality control (QC) on the dataset consisted of removing individuals with sex mismatch errors (eg. reported sex different from inferred sex) or had a sample call rate <90% and removing palindromic variants or variants with a call rate <95%. Genotyped data for unique samples were phased (using EAGLE) and imputed to the TOPMed Reference Panel (Freeze 5) on the TOPMed Imputation server. Eigenstrat Principal Components Analysis (PCA) was used to identify the genetic substructure of the entire PMBB population and we performed quantitative discriminant analysis (QDA) on all samples to determine their genetically informed ancestry. For this analyses, 1000 Genomes samples were used as training set with labels and PMBB samples were used as a testing set to determine their ancestry. Individuals from genetically-determined African and European ancestry were included in the analysis. The first ten ancestry-specific principal components for the African and European individuals were used as covariates to account for genetic ancestry^18^.

### Phenotyping of Penn Medicine Biobank participants

We evaluated 19,935 individuals in PMBB with available genotyping data who had passed genotype QC and had at least one physician encounter, who together had 4.9 million health care encounters. Cancer cases were identified from the EHR using International Classification of Diseases (ICD)-9 or -10 billing codes. In order to determine the number of instances of ICD9/10 codes needed to identify cancer cases, manual chart review was performed for 2365 individuals with at least one ICD9/10 billing code for prostate cancer. The positive predictive value (PPV) of one ICD9/10 prostate cancer billing code was 94% (**Figure S1**). Based on this, we used a similar strategy to identify individuals with bladder, breast, colorectal, endometrial, esophageal, glioma, lung, melanoma, oral cavity and pharynx, ovarian, pancreatic, prostate, renal, testicular, and thyroid cancer (**Table S1, Figure S2**). Controls were defined as individuals with no ICD-9/10 codes for invasive cancer, benign, in situ, or secondary neoplasms (Figure S2). The African genetic ancestry dataset contained 8,711 individuals and the European genetic ancestry dataset contained 9,788 individuals. Of these, 8,673 African and 9,759 European genetic ancestry individuals had complete genotype, phenotype, and covariate data.

### Single nucleotide polymorphism (SNP) Selection

Summary statistics were obtained using the most recent study available in the GWAS Catalog^19^ for each of the 15 cancers in this study (**Table S2**). SNPs were chosen based on a p-value threshold in the largest published GWAS of p<1×10^−6^. SNPs were pruned for linkage disequilibrium based on pairwise genotypic correlation at r^2^ 0.1 (**Table S3**) using PLINK 1.9^20^.

### Polygenic risk score generation and statistical analysis

PRS for each individual was calculated using PLINK 1.9 by summing the LD-pruned SNP variants and weighing their corresponding effect sizes using the odds ratios or beta values reported in the original GWAS study. We standardized each cancer PRS to mean of 0 and standard deviation (SD) of 1 and used logistic regression to test for the association between cancer polygenic risk score and cancer phenotype and to compare the top versus bottom quintiles of polygenic risk, controlling for age, sex, and the first ten within-ancestry principal components (PC) as covariates. We performed a subgroup analysis for cancer cases with > 100 individuals. Area under the curve (AUC) for primary phenotype was determined using the package pROC^21^. All statistical analyses were performed in R 4.0.3.

## RESULTS

Of the 9,759 individuals of European genetic ancestry (men= 6,379; women=3,380, mean age = 64.1) with complete information (genotype, phenotype and covariate), 1,761 (25.1%) had at least one ICD-9/10 code for at least one of the 15 different types of cancer (bladder, breast, colorectal, endometrial, esophageal, glioma, lung, melanoma, oral cavity, ovarian, pancreatic, prostate, renal, testicular, thyroid) (**Table 1**). There were 8,673 individuals of African genetic ancestry with complete information (men = 3,200, women =5,473, mean age = 51.9) in PMBB, and of these 1,532 (17.6%) had at least one ICD-9/10 code recorded for a cancer of interest (**Table 1**).

**Table 1:** Number of cancer cases and controls for each polygenic risk score cancer study.

| Cancer Type | Controls | European |  |  | Controls | African |  |  |
| --- | --- | --- | --- | --- | --- | --- | --- | --- |
|  |  | Cases | Males |  |  | Cases | Males |  |
|  |  |  | n | % |  |  | n | % |
| Bladder | 6466 | 176 | 159 | 90.3 | 6099 | 92 | 60 | 65.2 |
| Colorectal | 6466 | 147 | 101 | 68.7 | 6099 | 138 | 55 | 39.9 |
| Esophageal | 6466 | 41 | 35 | 85.4 | 6099 | 22 | 14 | 63.6 |
| Glioma | 6466 | 30 | 21 | 70.0 | 6099 | 15 | 8 | 53.3 |
| Lung | 6466 | 290 | 170 | 58.6 | 6099 | 204 | 74 | 36.3 |
| Melanoma | 6466 | 229 | 168 | 73.4 | 6099 | 16 | 8 | 50.0 |
| Oral Cavity and Pharynx | 6466 | 80 | 60 | 75.0 | 6099 | 31 | 23 | 74.2 |
| Pancreatic | 6466 | 47 | 35 | 74.5 | 6099 | 50 | 31 | 62.0 |
| Renal | 6466 | 182 | 149 | 81.9 | 6099 | 176 | 115 | 65.3 |
| Thyroid | 6466 | 124 | 66 | 53.2 | 6099 | 122 | 19 | 15.6 |
| Female-only cancer cases |  |  |  |  |  |  |  |  |
| Breast | 2293 | 210 |  |  | 3996 | 332 |  |  |
| Endometrial | 2293 | 48 |  |  | 3996 | 77 |  |  |
| Ovarian | 2293 | 36 |  |  | 3996 | 41 |  |  |
| Male-only cancer cases |  |  |  |  |  |  |  |  |
| Prostate | 4173 | 470 |  |  | 2103 | 443 |  |  |
| Testicular | 4173 | 18 |  |  | 2103 | 6 |  |  |

African: genetically determined African ancestry; European: genetically determined European ancestry

Based on publicly available summary statistics from genome-wide association studies, we explored the association of cancer PRS based on genome-wide significant SNPs with the burden of their corresponding cancer phenotype. Among the cancer cases >100 participants from individuals of European genetic ancestry, the PRS for breast (OR 1.29, 95% CI 1.12-1.49, p=0.0004), colorectal (OR 1.25 (95% CI 1.06 – 1.48, p= 0.007), melanoma (OR 1.39, 95% CI 1.23 – 1.58, p=3.1 x 10^−7^), and prostate cancer (OR 1.47, 95% CI 1.33 – 1.63, p=1.3 ×10^−13^) were significantly associated with their respective cancer phenotypes (**Figure 1a**). For the remaining cancers we could not detect a significant association between the PRS and their respective cancers (**Figure 1a, Table S41**). Compared to those in the lowest PRS quintile, individuals of European genetic ancestry in the top PRS quintile also had a 20% greater odds of colon (OR 1.21, 95% CI 1.02 – 1.44), nearly 30% greater odds of bladder (OR 1.29, 95% CI 1.07 – 1.55), breast (OR 1.28, 95% CI 1.09 – 1.50), and nearly 40% and 50% greater odds of melanoma (OR 1.36, 95% CI 1.17 – 1.59) and prostate cancer respectively (OR 1.45, 95% CI 1.29 – 1.63). (**Figure S3**).

**Figure 1:**
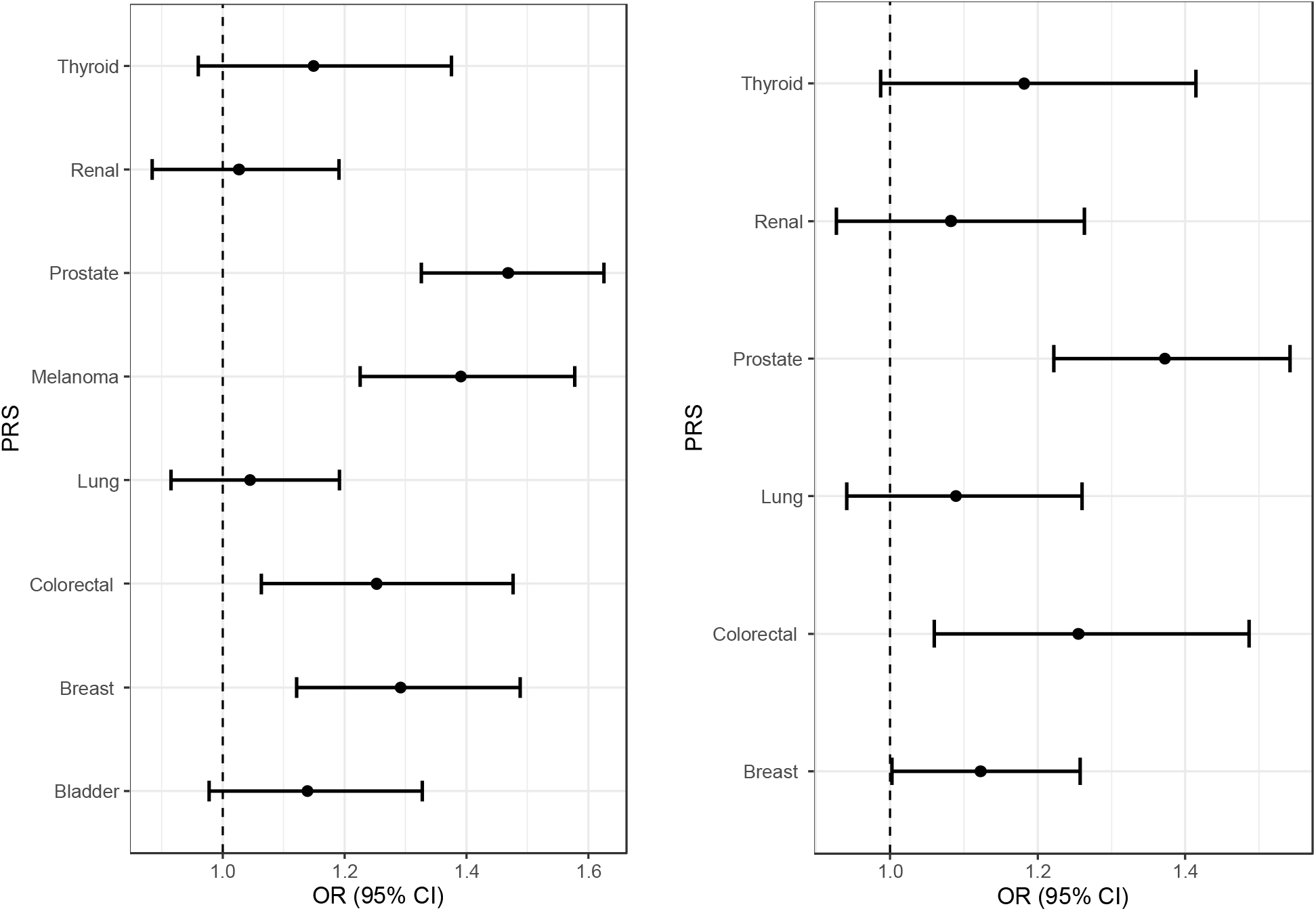
Association of PRS with cancer in academic biobank participants. **a**) Forest plots demonstrating the odds ratio for specific cancers of a PRS comparing the top and bottom of scores divided at the median for cancers with greater than 100 cases in individuals of genetically determined European ancestry (European; breast, bladder, colorectal, lung, melanoma, renal, prostate and thyroid). **b**) Forest plots demonstrating the odds ratio for specific cancers of a PRS comparing the top and bottom of scores divided at the median and comparing the top to the bottom quintile of scores for cancers with greater than 100 cases in individuals of genetically determined African ancestry (African; breast, colorectal, lung, renal, prostate and thyroid).

Among the cancer cases with African genetic ancestry > 100 individuals, the PRS for breast (OR 1.12, 95% CI 1.00 – 1.26, p=0.05), colon (OR 1.26, 95% CI 1.06 – 1.49, p=0.008), prostate (OR 1.37, 95% CI 1.22 – 1.54, p=9.7 ×10^−08^) were significantly associated with their respective cancer phenotypes (**Figure 1b**), and the PRS for thyroid was approaching significance (OR 1.18, 95% CI 0.99-1.41, p=0.07). Compared to individuals in the lowest PRS quintile, individuals of African genetic ancestry in the top PRS quintile also had nearly 30% greater odds of thyroid (OR 1.28, 95% CI 1.03 – 1.60), colorectal (OR 1.27, 95% CI 1.05 – 1.53), and nearly 40% greater odds of prostate cancer (OR 1.39, 95% CI 1.22 – 1.58) (**Figure S4**).

We examined the AUCs for a full logistic regression model incorporating age, sex, ancestry-specific principal components and PRS in the cancer PRS significantly associated with their phenotypes (breast, colorectal, melanoma, and prostate in European genetic ancestry; breast, colorectal and prostate in African genetic ancestry). Among the individuals of European genetic ancestry, the AUC ranged from 0.641 – 0.732 (**Table 2**). Overall, 90-97% of the AUC from the full model was explained by age alone (AUC for age alone: 0.580 – 0.707), which increased to 96-99% with the additional covariates of PCs and sex. The addition of the PRS explained 1-4% of the remaining AUC. On average, the inclusion of PRS improved the discriminatory accuracy in European ancestry individuals by over 2% compared with the model consisting of principal components (PCs), sex, and age alone, in breast, prostate and testicular cancers. Among individuals of African genetic ancestry, AUC for the model incorporating age, sex, principal components and statistically significant PRS ranged from 0.742 - 0.818. Among individuals of African genetic ancestry, age explained 98-99% of the AUC for the full model, which increased to 99-100% of the AUC for the full model with the addition of PCs and sex. The inclusion of PRS improved the discriminatory accuracy in individuals of African ancestry by a smaller amount than in individuals of European ancestry (AUC difference in African ancestry: PRS_breast_ - 0.002, PRS_prostate_ = 0.007). Overall, as compared to individuals of European genetic ancestry, in individuals of African genetic ancestry, the PRS contributed a smaller proportion of the AUC for the full model (average AUC difference 0.018 vs. 0.0005).

**Table 2:** Discriminatory accuracy of components of the PRS in individuals of genetically determined African and European ancestry in the PMBB.

| Cancer | European AUCs |  |  |  |  | African AUCs |  |  |  |  |
| --- | --- | --- | --- | --- | --- | --- | --- | --- | --- | --- |
|  | PRS | Age | PCs,<br>Age,<br>Sex | AUC - Full<br>Model | AUC<br>addition<br>PRS | PRS | Age | PCs<br>Age,<br>Sex | AUC - Full<br>Model | AUC<br>addition<br>PRS |
| Breast | 0.563 | 0.580 | 0.617 | 0.641 | 0.024 | 0.533 | 0.737 | 0.740 | 0.742 | 0.002 |
| Prostate | 0.599 | 0.707 | 0.709 | 0.732 | 0.023 | 0.563 | 0.809 | 0.811 | 0.818 | 0.006 |
| Testicular | 0.598 | 0.625 | 0.758 | 0.779 | 0.021 | 0.712 | 0.558 | 0.778 | 0.870 | 0.092 |
| Melanoma | 0.614 | 0.655 | 0.694 | 0.710 | 0.017 | 0.636 | 0.698 | 0.751 | 0.758 | 0.006 |
| Esophageal | 0.585 | 0.589 | 0.732 | 0.747 | 0.015 | 0.507 | 0.775 | 0.809 | 0.809 | 0.000 |
| Ovarian | 0.523 | 0.586 | 0.654 | 0.667 | 0.013 | 0.561 | 0.674 | 0.715 | 0.734 | 0.019 |
| CRC | 0.568 | 0.653 | 0.680 | 0.688 | 0.009 | 0.570 | 0.739 | 0.747 | 0.755 | 0.008 |
| Endometrial | 0.554 | 0.594 | 0.657 | 0.666 | 0.008 | 0.518 | 0.803 | 0.811 | 0.812 | 0.001 |
| Thyroid | 0.549 | 0.530 | 0.634 | 0.637 | 0.003 | 0.539 | 0.644 | 0.731 | 0.736 | 0.004 |
| Glioma | 0.551 | 0.566 | 0.675 | 0.678 | 0.003 | 0.495 | 0.648 | 0.715 | 0.713 | -0.002 |
| Bladder | 0.530 | 0.703 | 0.771 | 0.772 | 0.001 | 0.502 | 0.844 | 0.860 | 0.860 | 0.000 |
| Lung | 0.521 | 0.626 | 0.646 | 0.647 | 0.001 | 0.523 | 0.830 | 0.837 | 0.838 | 0.001 |
| Renal | 0.516 | 0.618 | 0.681 | 0.682 | 0.000 | 0.516 | 0.754 | 0.785 | 0.785 | 0.001 |
| Oral Cavity and<br>Pharynx | 0.521 | 0.611 | 0.669 | 0.668 | -0.001 | 0.530 | 0.793 | 0.843 | 0.846 | 0.003 |
| Pancreatic | 0.482 | 0.659 | 0.730 | 0.728 | -0.002 | 0.518 | 0.770 | 0.806 | 0.808 | 0.002 |

African: African ancestry; AUC: area under the curve; European: European ancestry; PC: principal components; PRS: polygenic risk score

## DISCUSSION

In this large retrospective case-control study, we evaluated the performance of PRS constructed using GWAS-identified cancer risk variants for 15 major cancers in a hospital-based biobank from a large academic medical center. Among individuals of European genetic ancestry, the PRS for breast, colorecal, melanoma, and prostate cancer were significantly associated with their respective cancer phenotypes. Among individuals of African genetic ancestry, the PRS for breast, colorectal, and prostate cancer were significantly associated with their cancer phenotype. For individuals of both European and African genetic ancestry, age contributed the highest proportion to the AUC (among 90-97% for European and 96-99% among African individuals) while the contribution of PRS was higher in individuals of European versus African genetic ancestry. Restricting to cancers with >100 cases, the average AUC difference was 0.018 in individuals of European compared to 0.0005 in individuals of African genetic ancestry.

Although the discriminatory ability of PRS to identify cancer patients versus controls was moderate as measured using AUCs, this is consistent with prior literature in the UK Biobank^22^. The PRS could further identify patients who are at higher risk of developing certain cancers in individuals of European genetic ancestry. For example in breast cancer, the further discriminatory ability of PRS in our prediction model is similar to addition of established clinical risk factors such as mammographic breast density^13^, BMI^23^, and family history^11^ to baseline breast cancer prediction models.

In contrast, the discriminatory ability of PRS in individuals of African genetic ancestry were extremely small in nearly all cancers tested in our study, consistent with attenuated performance of PRS in non-White breast cancer patients^24^. It is well known that three-quarters of all GWAS studies have been performed exclusively in individuals of European genetic ancestry^14^. The lack of variant representation, loci diversity, and linkage disequilibrium patterns between populations among individuals of African genetic ancestry likely explains the smaller improvement of the model prediction compared with the genetic principal components alone. We predict that as GWAS for cancers in populations of African genetic ancestry approach the size of those currently reported in populations of European genetic ancestry, it will be possible to construct African ancestry-specific PRS with a discriminative capability similar to what we currently see in European ancestry populations. Multi-ethnic PRS have shown promise in prostate cancer patients of African genetic ancestry^25^.

We acknowledge limitations to our analysis. First, our cancer-free group was obtained from an academic biobank at a tertiary care academic center, where patients may have higher comorbidities and not necessarily reflect the general population. Second, currently our full model contains age, sex, principal components, and PRS, but the precise predictive capability of PRS in combination with other previously validated clinical attributes, such as family history, BMI, and breast density, is unknown. Follow-up of this study population with additional clinical factors could improve the performance of each cancer PRS.

We showed that genetic liability for cancer is associated with its corresponding cancer phenotypes as classified by the electronic health record, with a more robust predictive capability of PRS in individuals of European versus African genetic ancestry. Expansion of GWAS in non-White populations is critically important to improve estimates of genetic risk, further tailoring PRSs as a reliable future tool for clinical cancer risk prediction across ancestry groups.

## Data Availability

The datasets generated during and/or analysed during the current study are available from the corresponding author on reasonable request.

## Data availability

Summary statistics used to create PRS for each of the 15 cancers are available from the GWAS Catalog, as detailed in Table S2. Individual-level data for the PMBB are not publicly available due to research participant privacy concerns; however, requests from accredited researchers for access to individual-level data relevant to this manuscript can be made by contacting the corresponding author.

## Financial Support

This study is funded by the NCI (K08CA215312, KNM), Burroughs Wellcome Foundation (#1017184, KNM), Basser Center for BRCA (KNM), Abramson Cancer Center (DM, KNM), Prostate Cancer Foundation (# 20YOUN02 KNM), The Jonathan and Plum Simons Precision Oncology Center of Excellence (KNM), NIAAA (K01AA028292, RLK), and the Veterans Association (IK2-CX001780, SMD). This publication does not represent the views of the Department of Veterans Affairs or the United States Government.

## Author Information

Conceptualization: L.W., H.D., R.K., K.M.; Data curation: : L.W., H.D., R.K., K.M.; Formal Analysis: H.D.; Funding acquisition: K.M.; Investigation: L.W., H.D., R.K., K.M.; Project Administration: L.W.; Supervision: R.K., K.M.; Writing – original draft: L.W.; Writing – review & editing: L.W., H.D., R.K., K.M., S.V., A.L., R.H., A.V., R.J., A.D., P.G., K.N., S.D., D.M., M.D.

## Ethics Declaration

All individuals who were recruited for the PMBB are patients of clinical practice sites of the University of Pennsylvania Health System. Appropriate consent was obtained from each participant regarding storage of biological specimens, genetic sequencing, access to all available EHR data and permission to recontact for future studies. The study was approved by the Institutional Review Board of the University of Pennsylvania. Website: https://www.itmat.upenn.edu/biobank/

## Conflict of Interest

SMD receives research support to the University of Pennsylvania from RenalytixAI and consulting fees from Calico Labs, both outside the current work. SMD is named as a co-inventor on a Government-owned US Patent application related to the use of genetic risk prediction for venous thromboembolic disease filed by the US Department of Veterans Affairs in accordance with Federal regulatory requirements. MDR is on the scientific advisory board for Goldfinch Bio and Cipherome. The remaining authors have no conflicts of interest to report.

## Regeneron Genetics Center Banner Author List and Contribution Statements

All authors/contributors are listed in alphabetical order. ***RGC Management and Leadership Team:*** Goncalo Abecasis, Ph.D., Aris Baras, M.D., Michael Cantor, M.D., Giovanni Coppola, M.D., Aris Economides, Ph.D., Luca A. Lotta, M.D., Ph.D., John D. Overton, Ph.D., Jeffrey G. Reid, Ph.D., Alan Shuldiner, M.D. Contribution: All authors contributed to securing funding, study design and oversight. All authors reviewed the final version of the manuscript. Other specific contributions are delineated below: ***Sequencing and Lab Operations:*** Christina Beechert, Caitlin Forsythe, M.S., Erin D. Fuller, Zhenhua Gu, M.S., Michael Lattari, Alexander Lopez, M.S., John D. Overton, Ph.D., Thomas D. Schleicher, M.S., Maria Sotiropoulos Padilla, M.S., Louis Widom, Sarah E. Wolf, M.S., Manasi Pradhan, M.S., Kia Manoochehri, Ricardo H. Ulloa. Contribution: C.B., C.F., A.L., and J.D.O. performed and are responsible for sample genotyping. C.B, C.F., E.D.F., M.L., M.S.P., L.W., S.E.W., A.L., and J.D.O. performed and are responsible for exome sequencing. T.D.S., Z.G., A.L., and J.D.O. conceived and are responsible for laboratory automation. M.P., K.M., R.U., and J.D.O are responsible for sample tracking and the library information management system. ***Genome Informatics:*** Xiaodong Bai, Ph.D., Suganthi Balasubramanian, Ph.D., Andrew Blumenfeld, Boris Boutkov, Ph.D., Gisu Eom, Lukas Habegger, Ph.D., Alicia Hawes, B.S., Shareef Khalid, Olga Krasheninina, M.S., Rouel Lanche, Adam J. Mansfield, B.A., Evan K. Maxwell, Ph.D., Mrunali Nafde, Sean O’Keeffe, M.S., Max Orelus, Razvan Panea, Ph.D., Tommy Polanco, B.A., Ayesha Rasool, M.S., Jeffrey G. Reid, Ph.D., William Salerno, Ph.D., Jeffrey C. Staples, Ph.D. Contribution: X.B., A.H., O.K., A.M., S.O., R.P., T.P., A.R., W.S. and J.G.R. performed and are responsible for the compute logistics, analysis and infrastructure needed to produce exome and genotype data. G.E., M.O., M.N. and J.G.R. provided compute infrastructure development and operational support. S.B., S.K., and J.G.R. provide variant and gene annotations and their functional interpretation of variants. E.M., J.S., R.L., B.B., A.B., L.H., J.G.R. conceived and are responsible for creating, developing, and deploying analysis platforms and computational methods for analyzing genomic data. ***Research Program Management:*** Marcus B. Jones, Ph.D., Jason Mighty, Ph.D., Lyndon J. Mitnaul, Ph.D. Contribution: All authors contributed to the management and coordination of all research activities, planning, and execution. All authors contributed to the review process for the final version of the manuscript.

## Notes

### Author Declarations

The Penn Medicine BioBank (PMBB) is a genomic and precision medicine cohort comprised of participants who actively consent for biospecimen collection and linkage of the their biospecimen to their electronic health record (EHR) data17. Participants were recruited between 2004-2020 to a University of Pennsylvania institutional review board approved study at the time of their medical appointment in the University of Pennsylvania Health System (n=60,232). Appropriate consent was obtained from each participant regarding storage of biological specimens, genetic sequencing, access to all available EHR data and permission to recontact for future studies. The study was approved by the Institutional Review Board of the University of Pennsylvania.

