## Supplementary Figures for "Performance of polygenic risk scores for cancer prediction in a racially diverse academic biobank"

### PPV of ICD9/10 codes for prostate cancer

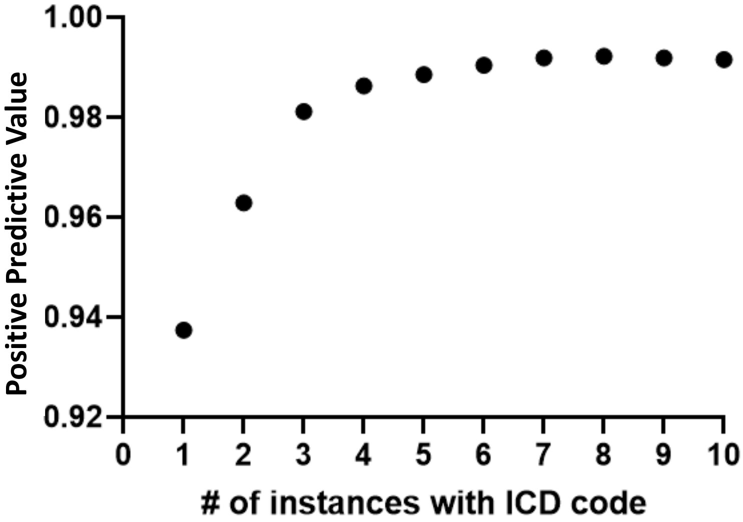


| # of ICD codes | 1 | 2 | 3 | 4 | 5 | 6 | 7 | 8 | 9 | 10 |
|---|---|---|---|---|---|---|---|---|---|---|
| Total | 2365 | 2129 | 1967 | 1897 | 1833 | 1775 | 1735 | 1681 | 1604 | 1540 |
| True positives | 2217 | 2050 | 1930 | 1871 | 1812 | 1758 | 1721 | 1668 | 1591 | 1527 |
| PPV | 0.937 | 0.963 | 0.981 | 0.986 | 0.989 | 0.990 | 0.992 | 0.992 | 0.992 | 0.992 |

**Supplementary Figure 1: Positive predictive value of ICD9/10 billing code based identification of cancer cases in PMBB.** Prostate cancer cases with the indicated number of instances of ICD9/10 codes for prostate cancer (C61 parent) and those in the cancer registry were manually reviewed to determine if truly had prostate cancer.

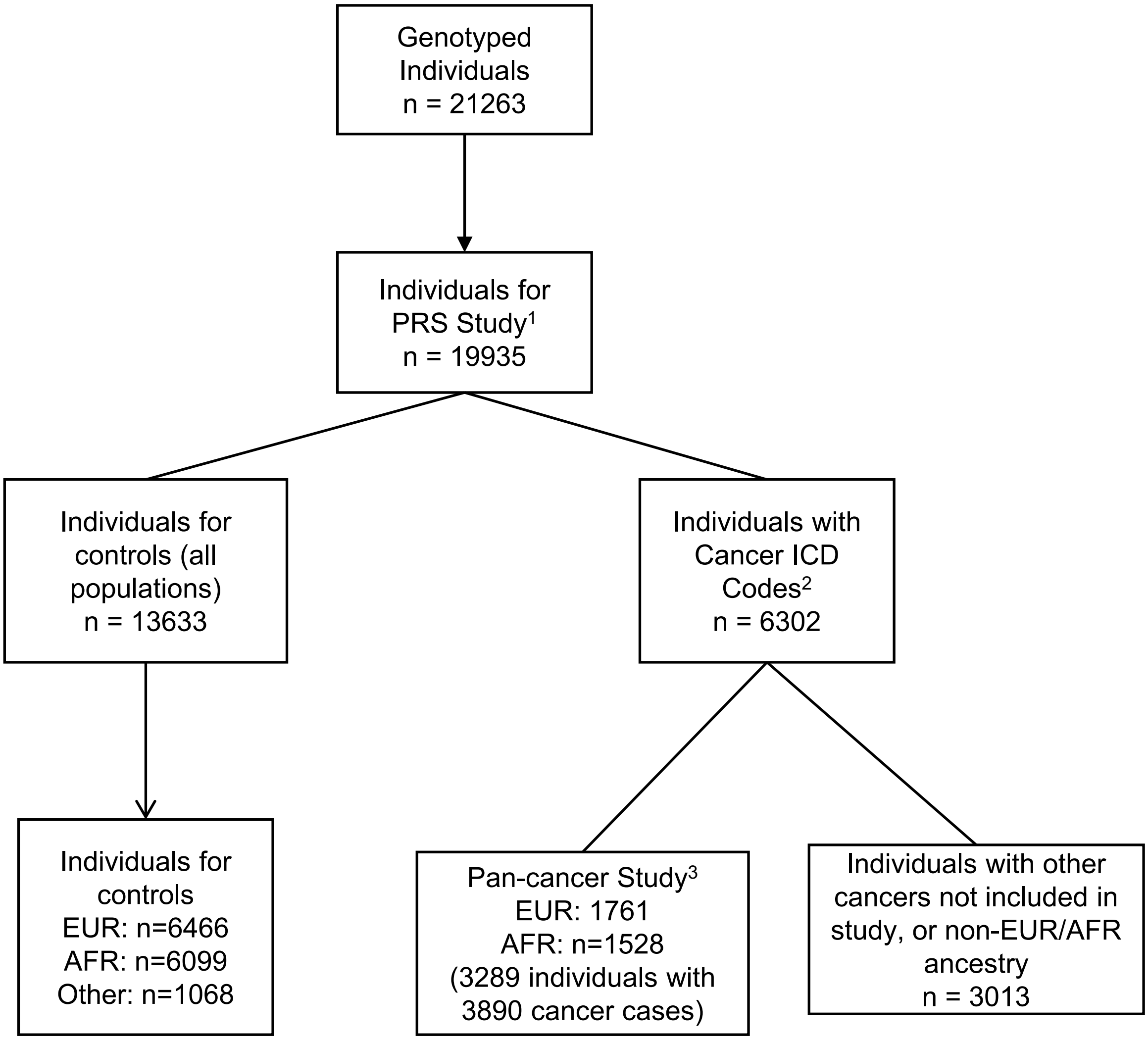


**Supplementary Figure 2: Cohort identification among genotyped individuals in PMBB.** From 21,263 individuals with SNP genotyping, 19,935 passed quality control measures. The number of instances of any ICD9/10 billing codes in the Electronic Health Record were obtained for each individual in the PMBB. Eigenstrat Principal Components Analysis (PCA) was used to identify the genetic substructure of the entire PMBB population and to assign genetically derived African ancestry (AFR), European ancestry (EUR), or Other ancestry to each individual. Cancer-free individuals had no ICD9/10 codes for any invasive cancer, nor any codes for benign, in situ or secondary neoplasms. Individuals with one or more cancers had at least one instance in the electronic health record of an ICD9/10 code corresponding to one or more of15 cancers with GWAS study summary statistics. Individuals with other cancers were excluded from the study.

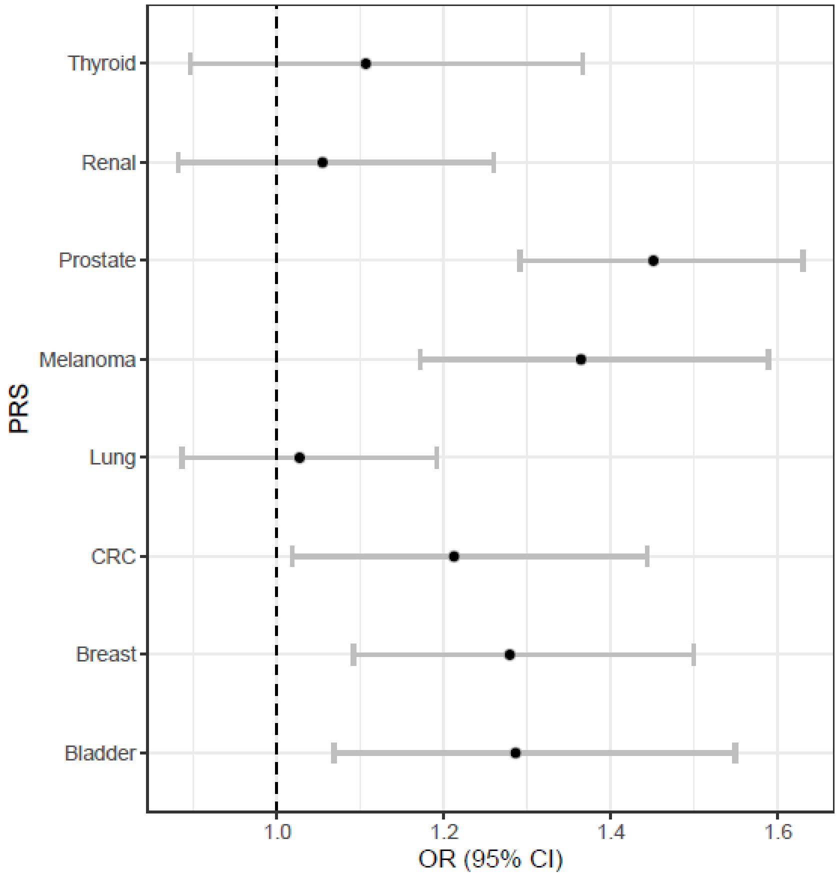


**Supplementary Figure 3: Association of PRS with cancer in EUR academic biobank participants.** Forest plots demonstrating the odds ratio for specific cancers of a PRS comparing the top to bottom quintile of scores for cancers with less than 100 cases in EUR (endometrial, esophageal, glioma, oral cavity, ovarian, pancreatic and testicular).

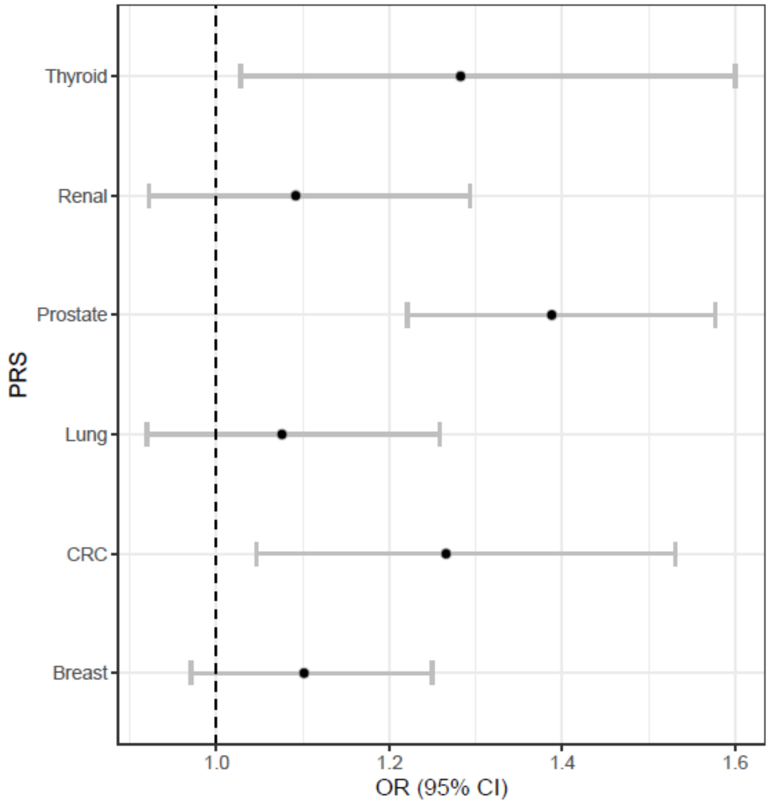


**Supplementary Figure 4: Association of PRS with cancer in AFR academic biobank participants.** Forest plots demonstrating the odds ratio for specific cancers of a PRS comparing the top and bottom of scores divided at the median and comparing the top to bottom quintile of scores for cancers with greater than 100 cases in AFR (bladder, endometrial, esophageal, glioma, melanoma, oral cavity, ovarian, pancreatic, testicular.
